# Fuzzy Autocatalytic analysis of Covid-19 outbreak in Malaysia

**DOI:** 10.1101/2020.05.17.20104968

**Authors:** Tahir Ahmad, Azmirul Ashaari, Siti Rahmah Awang, Siti Salwana Mamat, Wan Munirah Wan Mohamad, Amirul Aizad Ahmad Fuad, Nurfarhana Hassan

## Abstract

The objective of this research is to demonstrate a mathematical technique to analyze the Covid-19 outbreak, particularly with respect to Malaysia. The technique is able to accommodate scarcity, quantity, and availability of the data set. The obtained results can offer descriptive insight for reflecting and strategizing actions in combating the pandemic.

## 1. Introduction

The public panic and discomfort on the ongoing Covid-19 outbreak remind us of the history of the 1918 Spanish Flu pandemic, whereby over 50 million people died worldwide. It was a deadly pandemic, indeed. The ongoing outbreak of coronavirus disease 2019 (Covid-19) has claimed 105 952 lives worldwide as of 12 April 2020, 08:00 GMT, according to the World Health Organization (WHO) (https://www.who.int/emergencies/diseases/novel-coronavirus-2019). Since the first case of pneumonia of unknown cause detected in Wuhan reported to the WHO Country Office in China on 31 December 2019 and followed by its declaration as a Public Health Emergency by the international body on 30 January 2020, researchers, scientists, and mathematicians have been racing in their efforts to stop the potential devastating assault by the coronavirus.

These efforts include Zhou et al. [1] alerted the world the menace of the virus through their publication in Nature. However, the researchers did not employ any specific mathematical tools in their work. Hamzah et al. [2] utilized a system of ordinary differential equations for Susceptible-Exposed-Infected-Removed (SEIR) in their predictive modeling of the Covid-19 outbreak. Similarly, Lin et al. [3] adopted a system of ordinary differential equations that previously used to model the pandemic 1918 Spanish Flu for describing the current Covid-19 outbreak. Recently, Forster et al. [4] analyzed the coronavirus genomes using the phylogenetic network, a special type of graph that has been primarily used in archaeological studies.

There are three main problems with respect to the Covid-19 outbreak, namely, the scarcity, quantity, and availability of data that are essential to produce a good reliable mathematical model. This is due to the fact that the outbreak is about six months old since the first case was reported. Therefore, a mathematical technique must be flexible and robust enough to deal with such identified shortcomings is needed to model the outbreak. In this paper, a suitable mathematical method is proposed, namely a fuzzy autocatalytic set, which is able to accommodate such constraints to analyze the current pandemic.

## 2. Methods

Generally, a graph represents a relationship between objects. Objects are represented as vertices and the relations by edges. Formally, the definition of a graph is as follows

### Definition 1

(see [5]). A graph is a pair of sets (*V, E*) where *V* is the set of vertices, and *E* is the set of edges.

Furthermore, another way to represent a graph is by its adjacency matrix. The definition of an adjacency matrix for a graph is given in Definition 2 below.

### Definition 2

(see [5]). An adjacency matrix of graph *G*(*V, E*) with *n* vertices is an *n*×*n* matrix denoted by *A*(*a*_i_*_j_*), where *a_ij_ = 1* if *E* contains a directed edge (*j, i*). It is an arrow pointing from vertex *j* to vertex *i*, and *a_ij_* = 0 otherwise.

**Figure 1:**
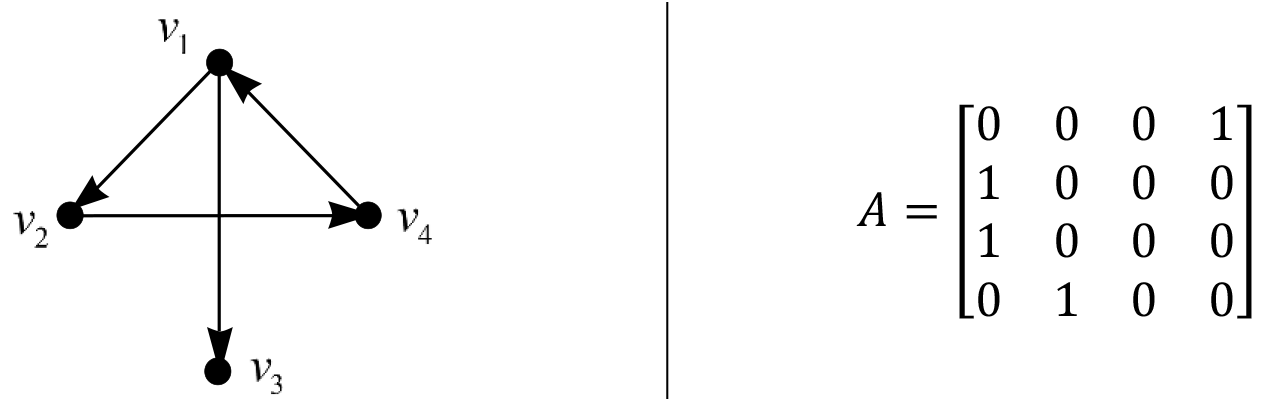
A directed graph with its adjacency matrix.

### 2.1 Fuzzy Autocatalytic Set

The concepts of graph and fuzzy set have given ‘birth’ to a new mathematical structure, namely, a fuzzy graph. Definition 3 indicates that vertices and edges are both fuzzy. In other words, the vertices and edges have values between 0 and 1. FIGURE 2 illustrates a fuzzy graph and its adjacency matrix.

#### Definition 3

(see [6]). A fuzzy graph *G*(*σ, μ*) is a pair of function *σ:S* → [0,1] and *μ:S* × *S* → [0,1] such that ∀*x,y* ∈ *S,μ*(*x,y*) ≤ *σ*(*x*) ∧*σ*(*y*).

An adjacency matrix of a fuzzy graph is defined as follows:

#### Definition 4

(see [6]). An adjacency matrix, *A* of a fuzzy graph *G* = (*V*, *σ, μ*) is an *n*×*n* matrix defined as *A =* (*a_ij_*) such that *a_ij_ = μ*(*ν_ij_, ν_i_*).

**Figure 2:**
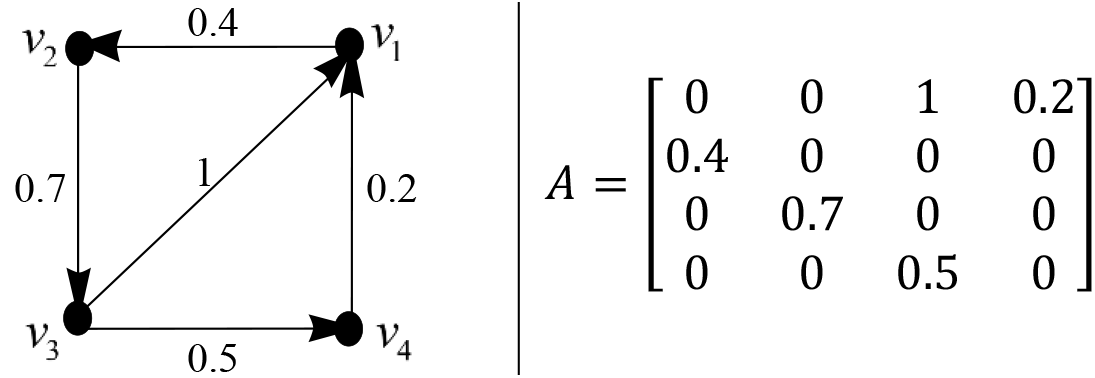
A fuzzy graph and its adjacency matrix.

The concept of autocatalysis was originated in chemistry, in particular, for the description of catalytic interaction between molecules [7], [8]. Further, Jain and Krishna [9] formalized the concept of an autocatalytic set (ACS) as a directed graph in which its vertices represent species and edges represent catalytic interactions among them. The formal definition of an ACS is given as follows.

#### Definition 5

(see [9]). An autocatalytic set is a subgraph, each of whose vertices has at least one incoming link from vertices belonging to the same subgraph.

Some examples of ACSs are illustrated in FIGURE 3. The simplest ACS is a vertex with 1-cycle.

**Figure 3:**
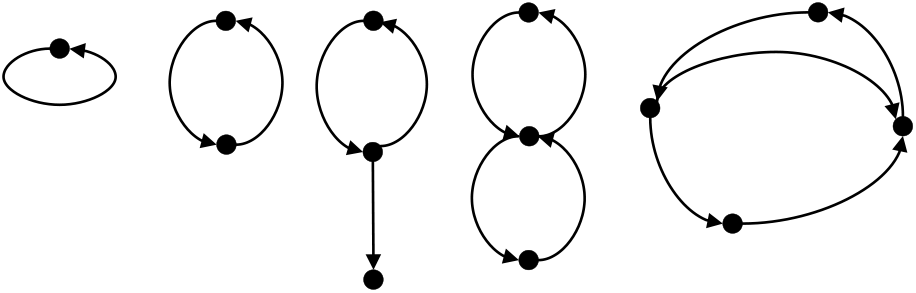
Some examples of ACS

The merger of the fuzzy graph and autocatalytic set has led to the idea of the fuzzy autocatalytic set (FACS) by Ahmad et al. [10]. The concept of FACS Covid-19 outbreak in Malaysia is depicted in FIGURE 4. The formal definition of FACS is laid as follows.

**Figure 4:**
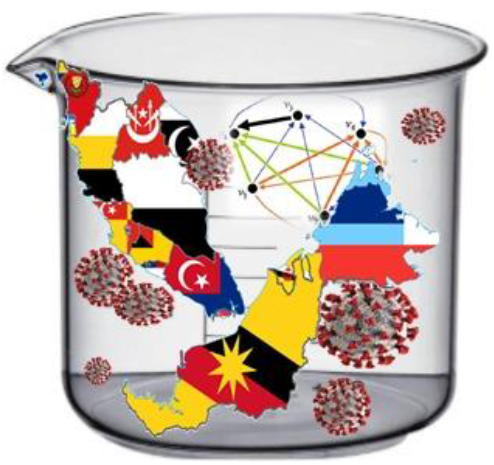
Fuzzy Autocatalytic Set of Covid-19 outbreak in Malaysia.

*Definition 6* (see [10]). A fuzzy autocatalytic set is a subgraph each of whose vertices has at least one incoming link with membership value, *μ*(*e_i_*) ∈ (0,1], *∀e_i_* ∈*E* from any other vertices are belonging to the same subgraph.

### 2.2 Dynamics of FACS

The adjacency matrix in FIGURE 1(b) and FIGURE 2(b) are then processed by the procedure outlined in [10], [11] and improved by [12], respectively. The outcomes of the process are determined via the following steps.

Step 1: Keeping *C*(*s* × *s)* matrix fixed, *x* evolved according to the following equation.

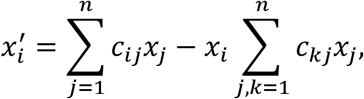

for time *t*, which is large enough for *x* to get reasonably close to its attractor ***X*** (Perron Frobenius Eigenvector). We denoted *X_i_* = *x_i_* (*t*).
Step 2: The set *L* of nodes *i* with the least value of *X_i_* is determined, i.e.

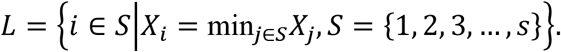 This is the set of “least fit” nodes, identifying the relative concentration of a variable in the attractor (or, more specifically, at *t*) with its “fitness” in the environment defined by the graph. The least fit node is removed from the system along with its links, leading a graph of *s* − 1 variables.
Step 3: *C* is now reduced to (*s* − 1) × (*s* − 1) matrix. The remaining nodes and links of *C* remained unchanged. All these *x_i_*(0 ≤ *x_i_* ≤ 1) are rescaled to keep

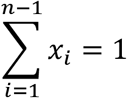

Repeat all the steps until the 2×2 matrix is attained.

FIGURE 5 illustrates the initial step (Step 1). Then one of the nodes with the least eigenvector is removed from the graph (Step 2). The node is removed along with its links, and the graph is left with a reduced number of nodes and links (Step 3). This process is then repeated until a graph with at least two nodes is attained.

**Figure 5:**
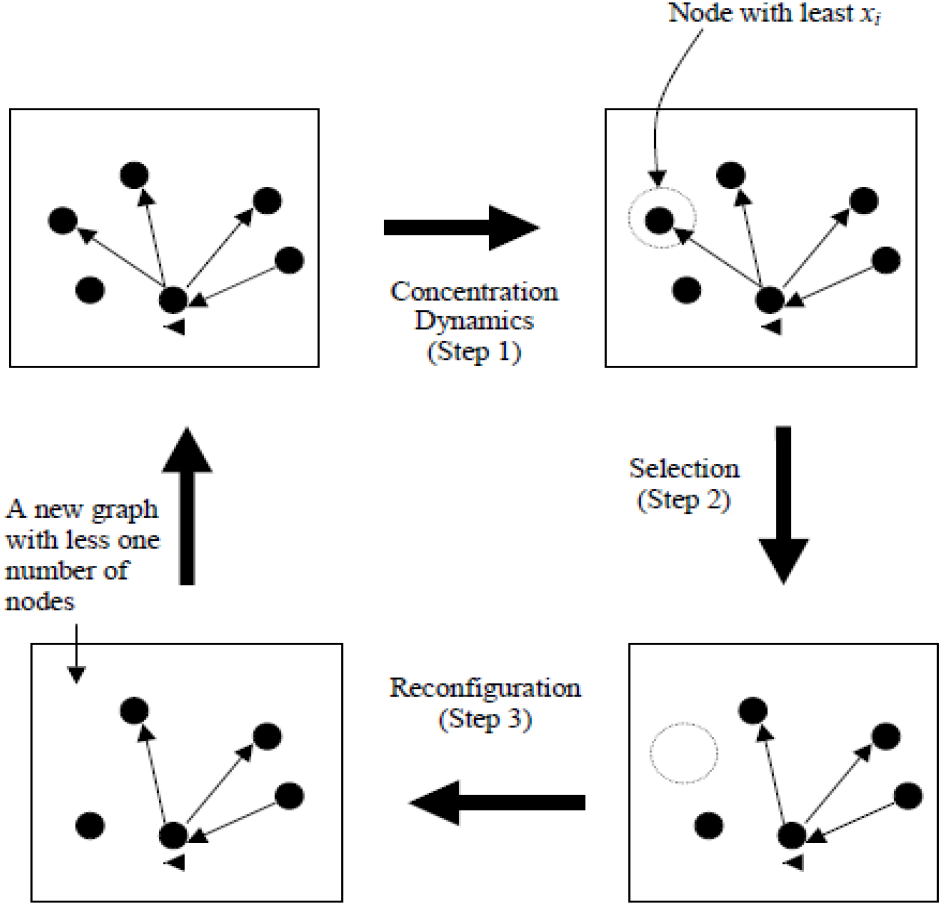
Schematic portrayal of the graph dynamics.

The procedure to transform the graph into 2D-Euclidean space is adopted from [13], which is based on the Laplacian matrix and solving a unique one-dimensional optimization problem in order to determine their coordinates. The general overview of the transformation is depicted in the following FIGURE 6.

**Figure 6:**
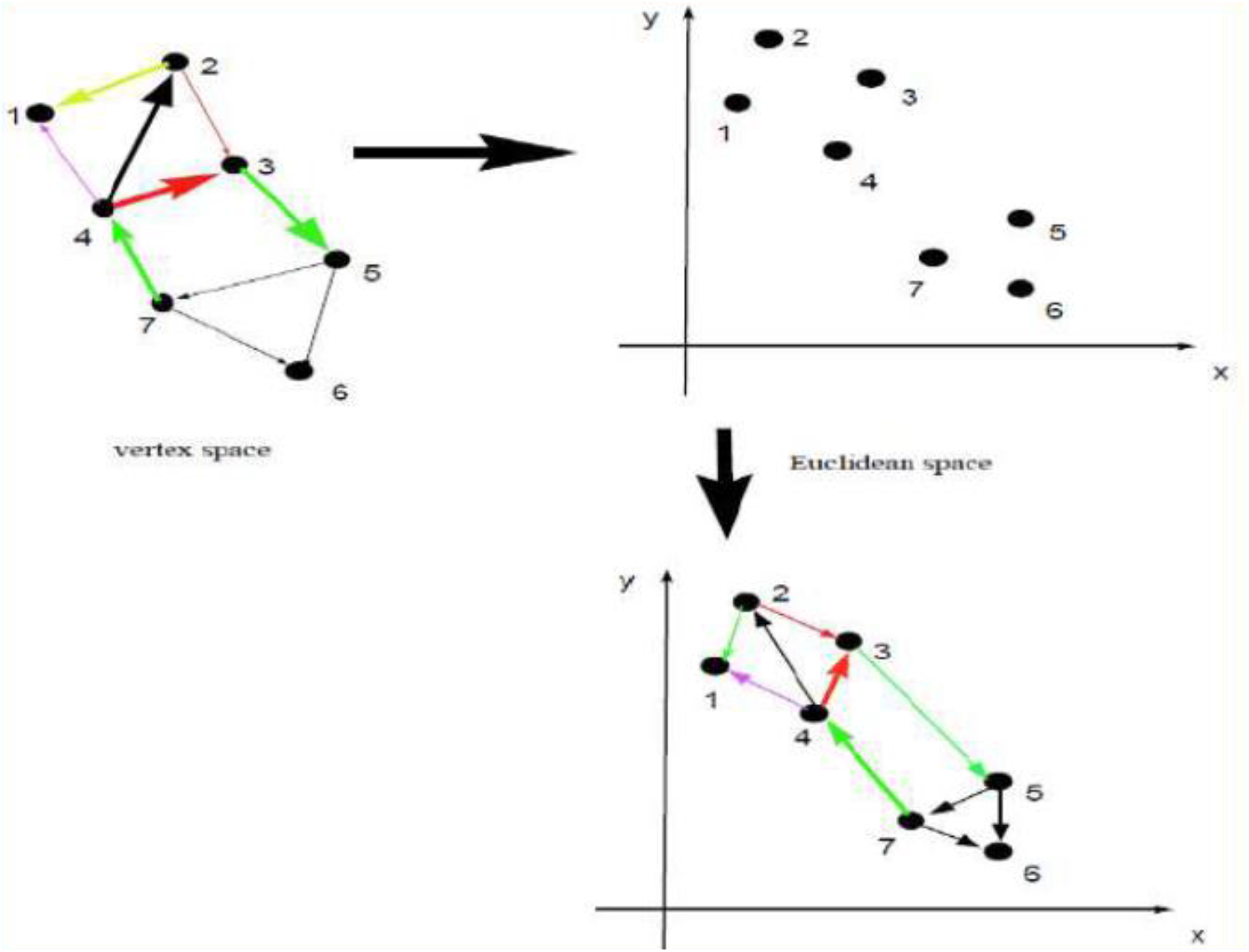
Schematic illustration transformation of the graph from vertex space to Euclidean space

## 3. Implementation

The technique described in Section 2 is implemented on two sets of data; Malaysia and its neighboring countries and states in Malaysia.

### 3.1 Malaysia and ist Neighboring Countries

The daily new reported cases of Covid-19 for Malaysia, Singapore, Thailand, Indonesia, and Brunei are obtained (publicly available) from European Centre for Disease Prevention and Control’s website (see https://www.ecdc.europa.eu/en/publicationsdata) from 1 February 2020 until 27 March 2020 (see FIGURE 7). In order to determine the pandemic signature of Covid-19 for these countries, we sampled the data from 12 to 27 March only. This is due to the fact that the plotted lines are clearly erratic for the sampled countries (refer FIGURE 7 and FIGURE 8) during that period.

**Figure 7:**
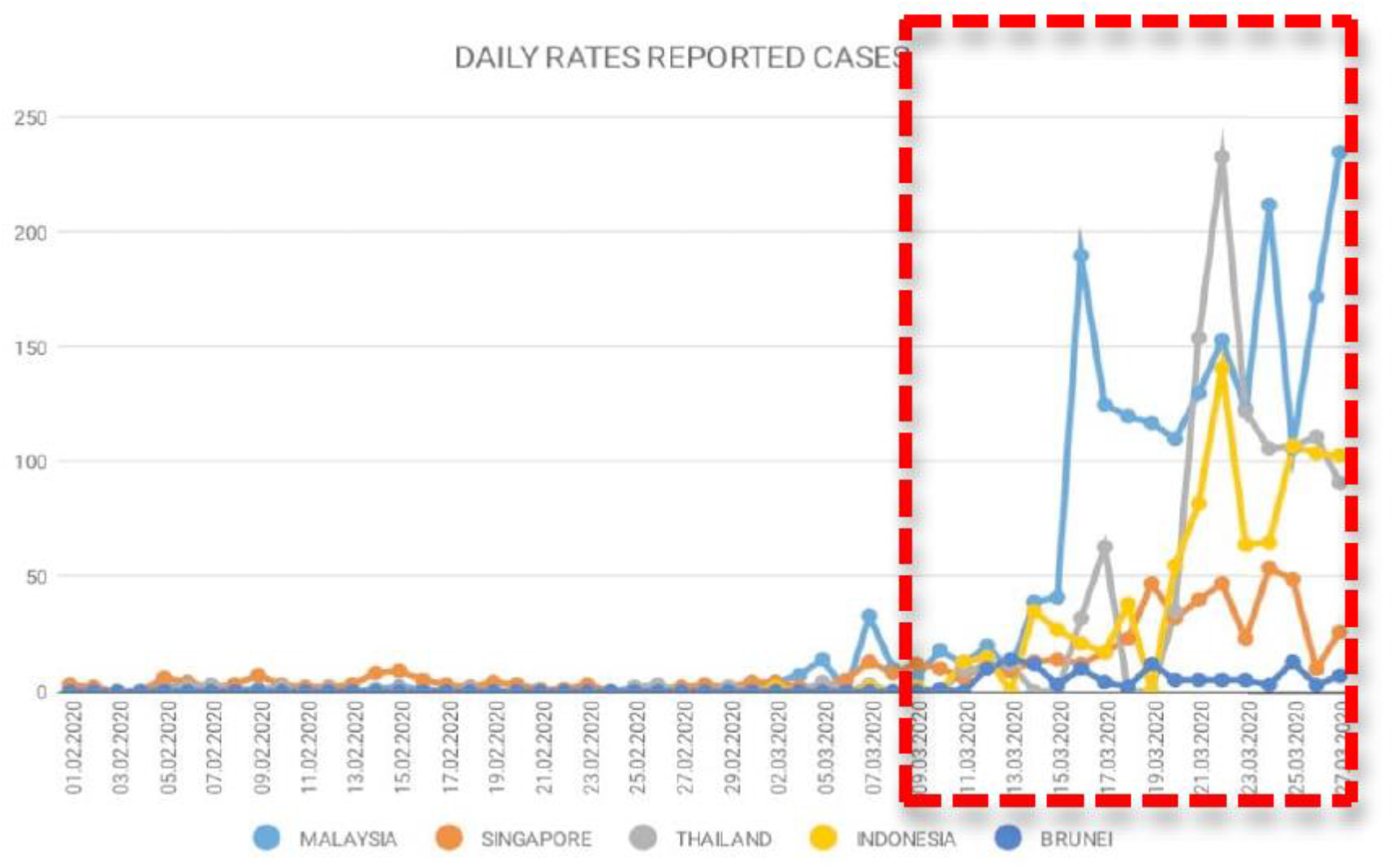
New cases with respect to Malaysia and its neighbors from 12 March to 27 March 2020.

**Figure 8:**
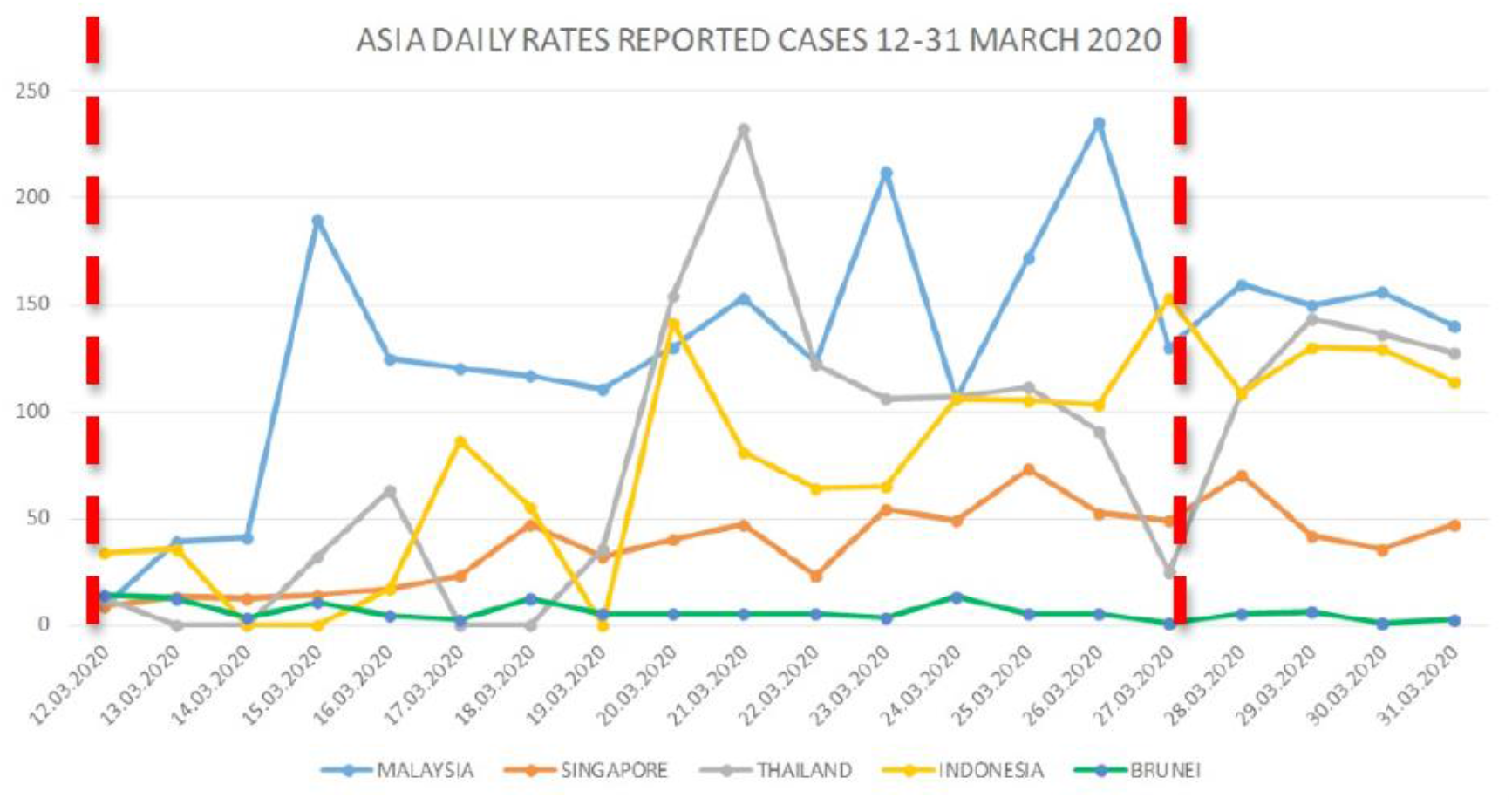
New cases with respect to Malaysia and its neighbors from 12 March to 27 March 2020

### 3.2 States of Malaysia

A set of data from 10 March to 10 April is obtained from the Ministry of Health, Malaysia (www.moh.gov.my), and presented in FIGURE 9.

**Figure 9:**
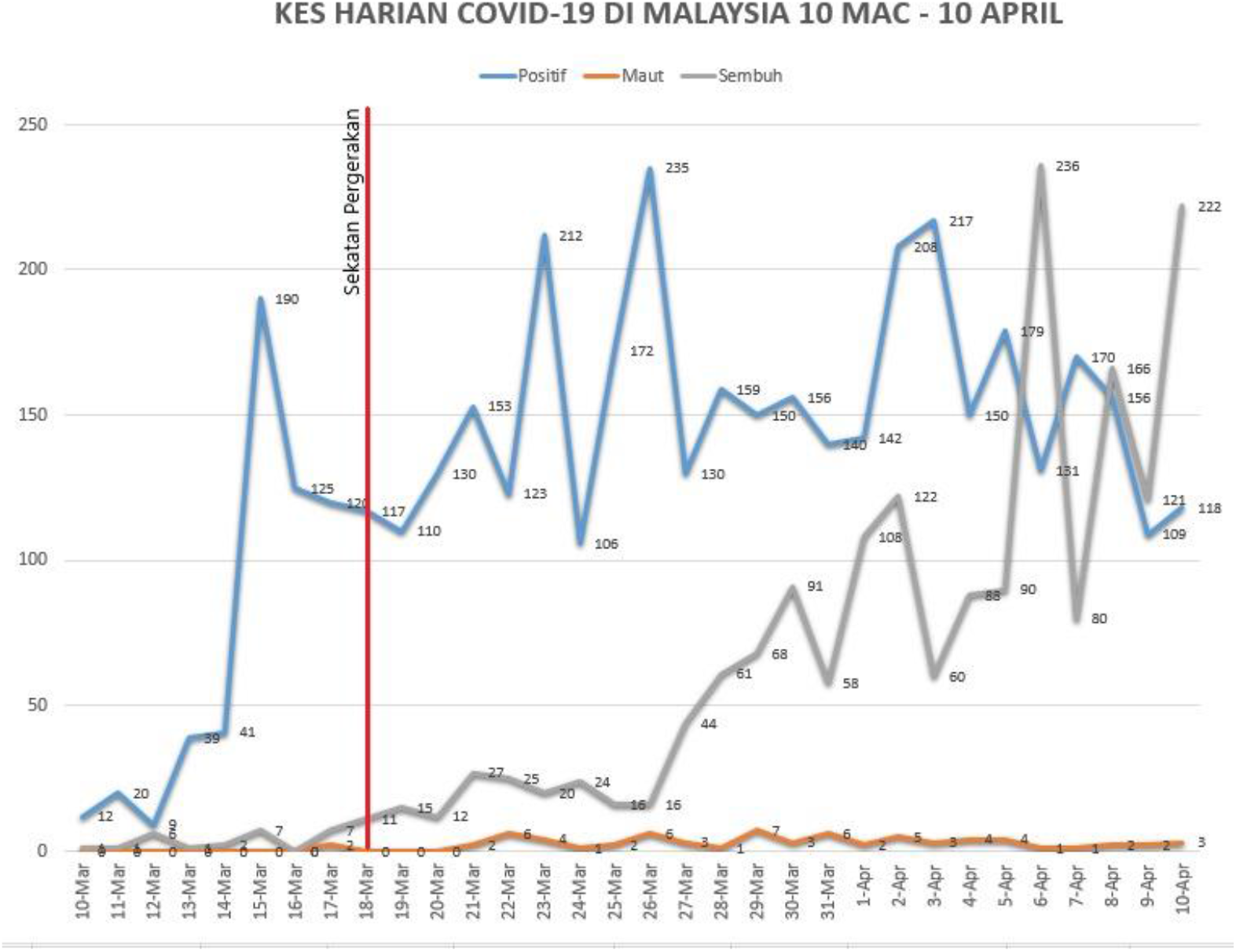
New cases from 10 March to 10 April 2020.

The breakdown of reported new cases between states in Malaysia from 28 March to 5 April is considered in our study (refer to FIGURE 10). The period is selected due to the erraticness of the data, as depicted in FIGURE 9 earlier.

**Figure 10:**
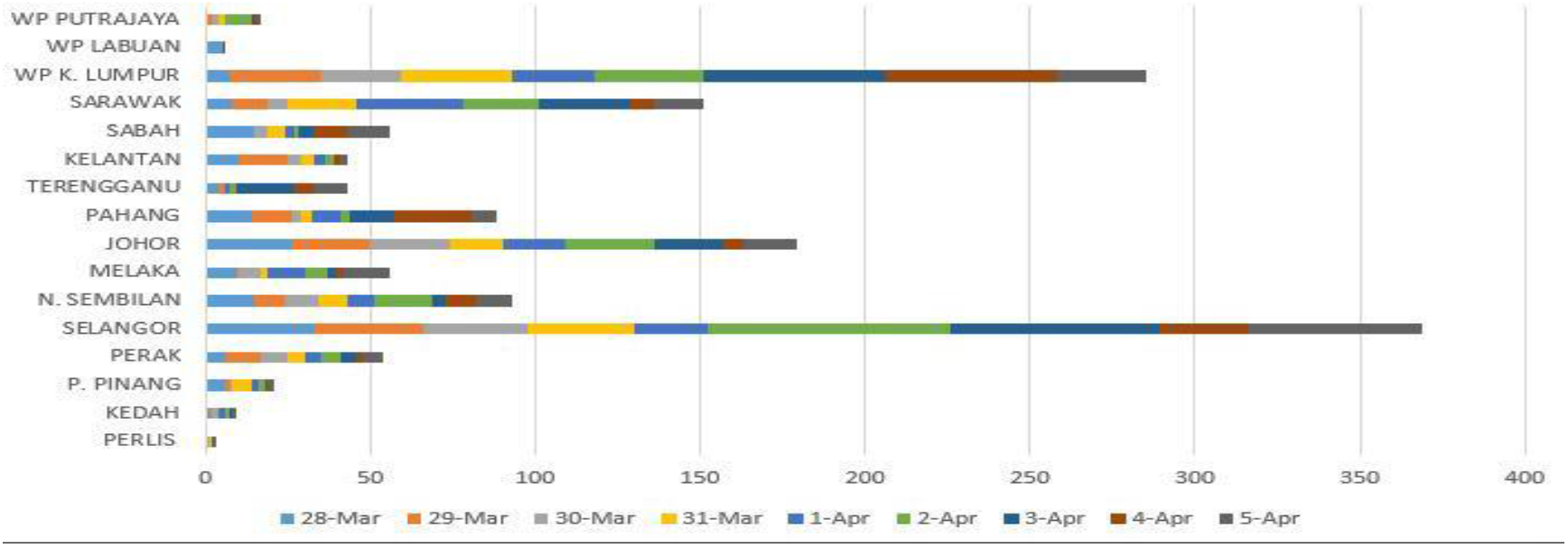
New cases in states of Malaysia from 28 March to 5 April 2020.

## 4. Analysis

Similarly, the fuzzy analysis of the graphs is divided into two identified demographic areas and presented in the following subsections.

### 4.1 Malaysia and its Neighbouring Countries

The graph and its adjacent matrix for data from 12 to 27 March are constructed and exhibited in FIGURE 11(a) and FIGURE 11(b), respectively. There are 16 days during the identified erratic interval. Hence the graph contains 16 vertices altogether. The impurities are considered as another vertex whereby it represents unidentified or undiscovered or unreported cases during the erratic period. This particular vertex acts as a buffer

**Figure 11:**
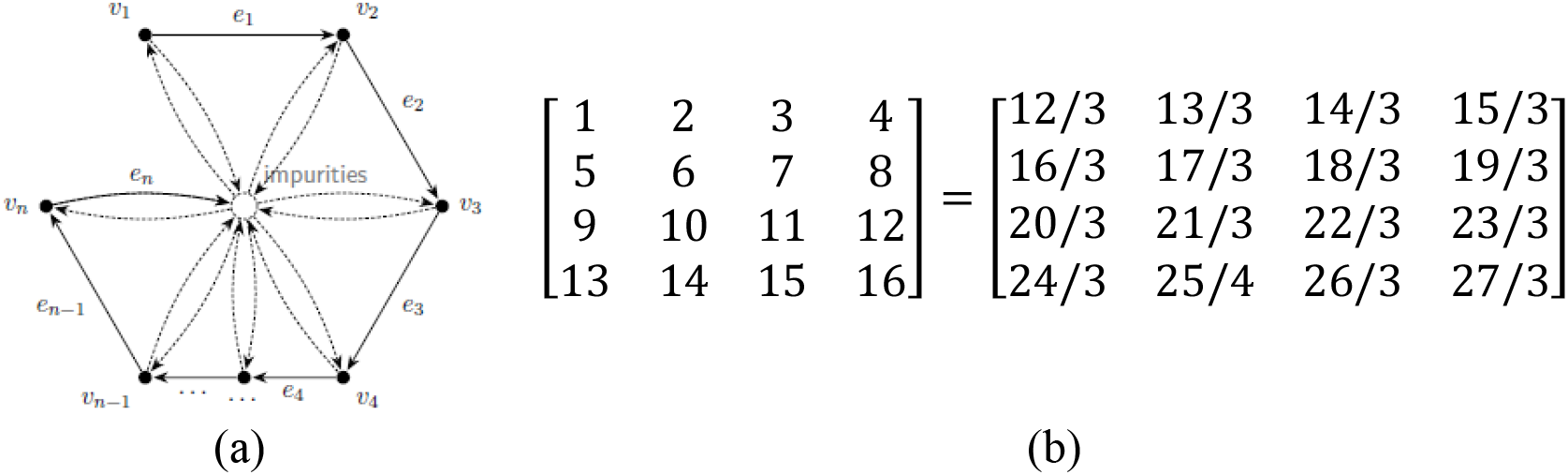
(a) Graph *C*with its (b) adjacency matrix for data from 12 March to 27 March 2020.

The dominance of each country (outcome) is identified for each day of the erratic interval and presented in the following TABLE 1.

**Table 1:** Output Matrix.

|  | **Results: $2 \times 2$ Variables Output Matrix (Represent dominant dates)** |
| --- | --- |
| **System 1 (Malaysia)** | $\begin{bmatrix} 11 & 12 \\ 15 & 16 \end{bmatrix} = 22/3, 23/3, 26/3, 27/3,$ |
| **System 2 (Singapore)** | $\begin{bmatrix} 11 & 12 \\ 15 & 16 \end{bmatrix} = 22/3, 23/3, 26/3, 27/3,$ |
| **System 3 (Thailand)** | $\begin{bmatrix} 11 & 12 \\ 15 & 16 \end{bmatrix} = 22/3, 23/3, 26/3, 27/3,$ |
| **System 4 (Indonesia)** | $\begin{bmatrix} 11 & 12 \\ 15 & 16 \end{bmatrix} = 22/3, 23/3, 26/3, 27/3,$ |
| **System 5 (Brunei)** | $\begin{bmatrix} 1 & 4 \\ 13 & 16 \end{bmatrix} = 12/3, 15/3, 24/3, 27/3,$ |

Malaysia, Singapore, Thailand, and Indonesia dominate on 03/22/2020, 03/23/2020, 03/26/2020, and 03/27/2020. This simply indicates that the outbreak has been spreading among the four countries with average daily reported cases of more than 60. On the other hand, Brunei recorded less than 10 cases daily during the erratic period.

Furthermore, the transformed graph into the 2D-Euclidean space reveals the nodes for each country are dispersed and scattered (see FIGURE 12: Clusters of assigned countries). However, Malaysia (S1) and Singapore (S2) nodes are closer to each other. This characteristic hint that daily reported cases for these two countries are quite similar, followed by Thailand (S3) and Indonesia (S4), whereas Brunei (S5) is isolated from the rest.

**Figure 12:**
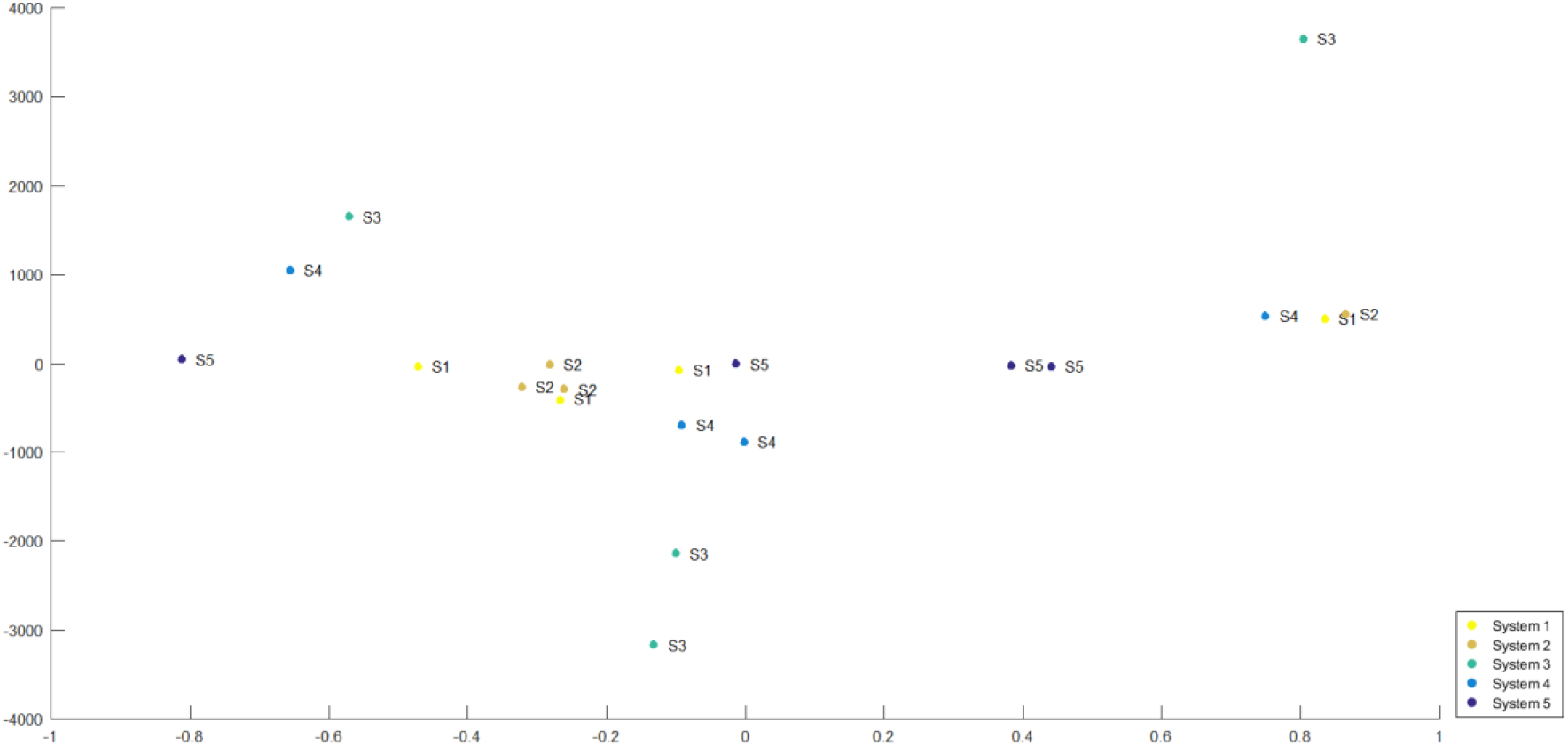
Clusters of assigned countries

To justify the obtained clusters, an ordinary graph for a daily rate of reported new cases over populations for each country is plotted in FIGURE 13: The daily rate of new cases over the population from 12 March to 31 March 2020. Clearly, the gap between lines for Malaysia and Singapore are closed, similarly Thailand and Indonesia. The line that represents Brunei is quite erratic. Nevertheless, the line represents Brunei seems stable after 24 March due to the country lockdown measure taken with respect to its border.

**Figure 13:**
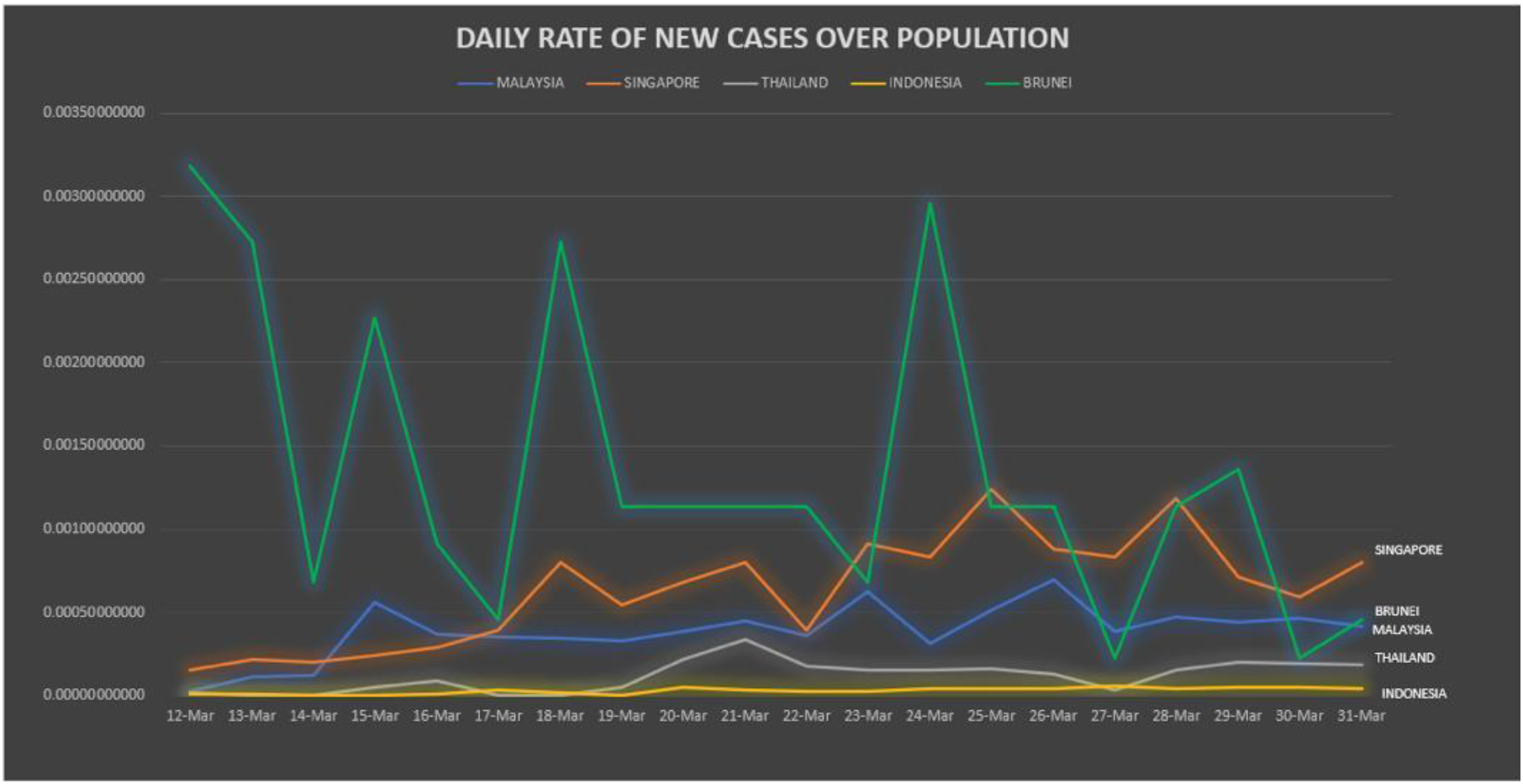
The daily rate of new cases over the population from 12 March to 31 March 2020.

### 4.2 States of Malaysia

Using FACS for sampled data 28 March to 5 April 2020, 16 states are identified and clustered, namely, **Cluster 1** contains Perlis, Kedah, Pulau Pinang and Perak. **Cluster 2** includes Selangor, Negeri Sembilan, Melaka and Johor whereas **Cluster 3** is made of Pahang, Trengganu, Kelantan and Sabah. Finally, Sarawak, WP Kuala Lumpur, WP Labuan and WP Putrajaya formed **Cluster 4**.

These clusters are then classified into three zones; zones 1, 2, and 3. Zone 1 is named as an **Under Control Zone** that comprises of Perlis, Kedah, Pulau Pinang, and Perak. These 4 states are scattered in Zone 1 (see FIGURE 14: Two phases of FACS clustering for states in Malaysia from 28 March to 5 April 2020.) that reflects their distinctness with low reported new cases.

**Figure 14:**
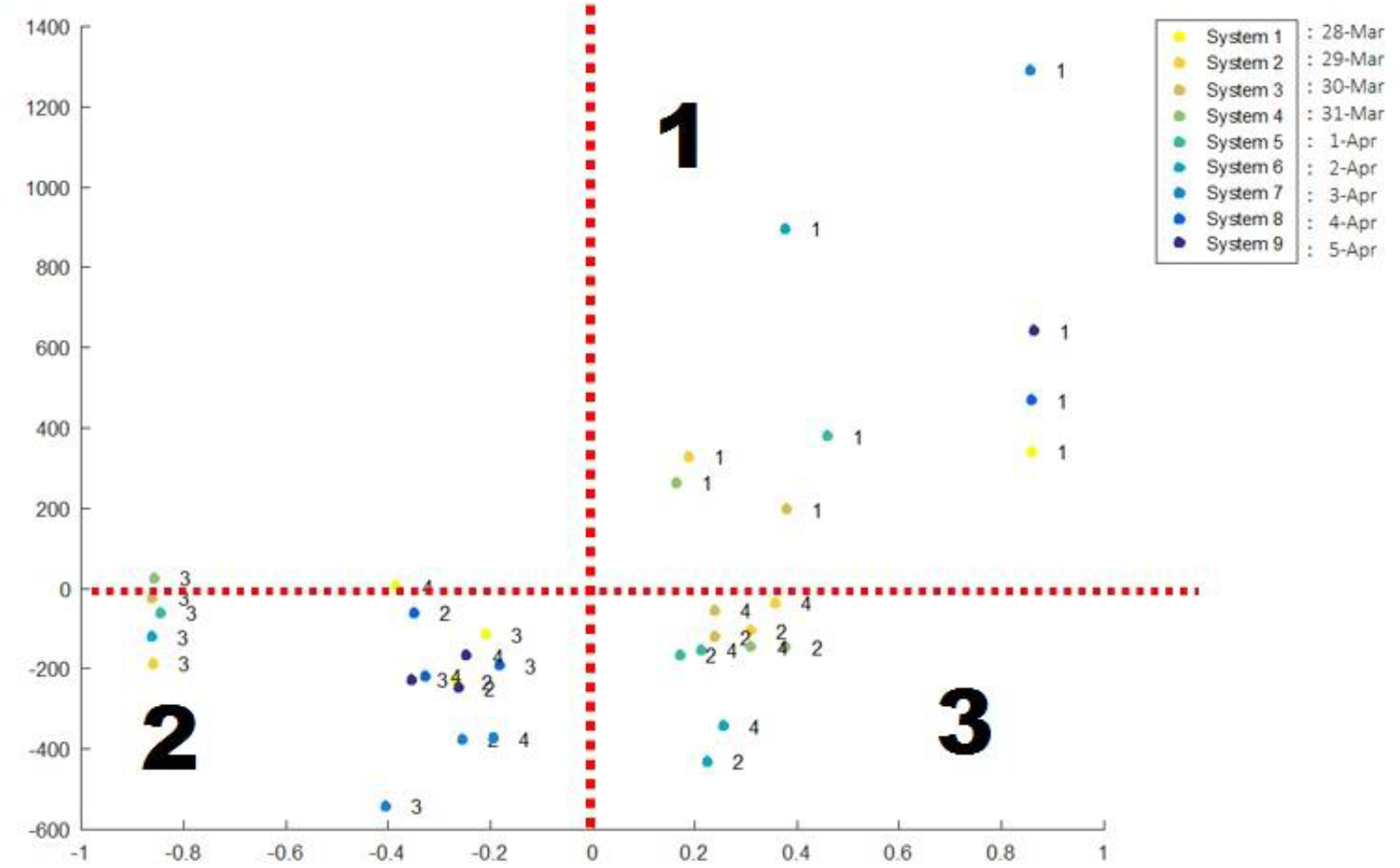
Two phases of FACS clustering for states in Malaysia from 28 March to 5 April 2020.

Zone 2 in the **Medium Zone** which consists of Pahang, Terengganu, Kelantan, and Sabah. Reported on increased new cases in these particular states only happened after 31 March. Even though Zone 2 is dominated by Cluster 3 but it is not total domination since there were a couple of instances where Cluster 2 and 4 popped up in the zone. Hence, the government has to pay attention to states in Cluster 3 because these states have the potential to move into Zone 3. On top of that, Zone 2 is clearly closed adjacent to Zone 3.

Zon 3 is the **Danger Zone** that is totally dominated by states in the west and south of Malaysia, such as Selangor, Negeri Sembilan, Melaka, Johor, WP Kuala Lumpur, WP Putrajaya, including Sarawak and WP Labuan. In fact, the government has gazetted 23 districts in these states as the red zone, namely, Putrajaya, Jasin, Negeri Sembilan, Hulu Langat, Petaling, Johor Bahru, Kuching, and Tawau. The district of Hulu Selangor in the state of Selangor has announced another red zone on 10 April. Our FACS analysis (refer to FIGURE 14) concurred with the list of states in red zones released by Crisis Preparedness and Response Centre (CPRC), Ministry of Health of Malaysia. Furthermore, we have predicted that WP Putrajaya is in Zone 3 with respect to data up to 5 April. True enough, WP Putrajaya was announced in the danger zone on 6 April by the government.

## 5. Conclusions

In this paper, we demonstrated a fuzzy autocatalytic analysis for the Covid-19 outbreak associated with Malaysia. The method is able to identify some significant features of the pandemic outbreak as well as some important predictions. The method can be used to model any future pandemic.

## Data Availability

The data are obtained from the Ministry of Health (MOH) Malaysia and National Institutes of Health Malaysia (NIH) (publicly available).

## Acknowledgments

We gratefully acknowledge the Ministry of Health (MOH) Malaysia and National Institutes of Health Malaysia (NIH) for allowing us to use their published data (publicly available). We thank the Faculty of Science and Azman Hashim International Business School, Universiti Teknologi Malaysia, for its tremendous support for this work.

